# Artificial Intelligence Analysis of ECG to Determine Fractional Flow Reserve (FFR)

**DOI:** 10.1101/2024.08.27.24312672

**Authors:** Łukasz Kalińczuk, Kamil Zieliński, Karol Artur Sadowski, Michael Leasure, Adam Butchy, Utkars Jain, Veronica A Covalesky, Rafał Wolny, Marcin Demkow, Maksymilian Opolski, Gary S Mintz

**Author notes:** Corresponding author: Łukasz Kalińczuk, MD, PhD Institute of Cardiology ul. Alpejska 42, 04–628 Warszawa, Poland /. equal senior authors (Maksymilian Opolski, Gary S Mintz). Sponsor: Heart Input Output Inc., Pittsburgh PA. Disclosures: ML, AB, UJ are owners and founders of Heart Input Output Inc.

## Abstract

**Background:** The current gold standard of coronary artery disease (CAD) diagnosis is invasive angiography, during which fractional flow reserve (FFR) measurement may be performed to confirm the clinical significance of a stenosis. The yield of routine and indiscriminate FFR in identifying hemodynamically significant stenoses is low. To combat this, we have developed an artificial intelligence model - ECGio – designed to be deployed at the point of care to determine FFR through the analysis of a resting digital 12-lead electrocardiogram (ECG), a fast, real-time, cost-effective, widely accessible, and safe diagnostic method.

This study assessed the ability of ECGio to train, tune, and test itself through a cross-validation paradigm to predict the presence of a reduced FFR in the left anterior descending artery in a patient population presenting for invasive FFR.

**Methods:** In a single-center study the ECGs of 209 consecutive patients (61.3±9.5 years, 35.4% female) from 2014 to 2021 were recorded within 7 days prior to angiography during which FFR was measured in the left anterior descending artery. Collected ECGs were used to train and test the AI model using a five-fold cross-validation methodology.

**Results:** The ability of ECGio to predict the presence of a reduced FFR (<0.80) in this cohort was a sensitivity, specificity, PPV, NPV, Accuracy, and F-1 Score of 43.2%, 86.7%, 64.0%, 73.6%, 71.3%, and 51.6%, respectively.

**Conclusions:** This study demonstrated the feasibility of using a deep learning AI algorithm to analyze a digital 12-lead ECG to provide a similar level of information as the invasive FFR.

**Highlights:**

- Coronary angiography is invasive and expensive and exposes the patient to radioactive dyes and risk of complications. Clinicians tend to fail-safe, overperforming testing and struggling to identify patients who would benefit from invasive testing resulting in procedures having low yield.
- Our AI model determines FFR by analyzing the patient’s resting digital 12 lead ECG which is fast, cheap, safe, and real-time.
- AI ECG analysis has the potential to play a crucial role in CAD diagnostics.

**Perspectives:** A major obstacle in CAD screening is that there is no quick, accurate, non-invasive test to differentiate patients that require additional testing and treatment from those that can be safely dismissed. The 12-lead digital ECG is the most easily acquired diagnostic test; it does not involve stress, radioactive dyes, or risk. Invasive FFR is the most accurate technique to identify an ischemia-producing stenosis. The current study demonstrated the feasibility of training AI to analyze ECG signals to recognize reduced FFR and even estimate the actual FFR value. Future studies will analyze patients who enter the diagnostic process through other clinical pathways in order to better understand the performance of ECGio in a more general patient population.

**Ethics:** This study was conducted in a deidentified, retrospective fashion from a pre-existing registry. The ethics committee waved the need for informed consent.

## Introduction

The current gold standard of coronary artery disease (CAD) diagnosis is invasive coronary angiography (1), during which fractional flow reserve (FFR) measurement may be performed to confirm the clinical significance of one or more stenoses and the need for revascularization. FFR is performed by measuring blood pressure both upstream and downstream to a stenosis to calculate the ratio at maximum hyperemia. The threshold of 0.80 distinguishes clinically significant lesions (≤0.80) from clinically insignificant ones (>0.80) (2). However, this test often results in a low yield of hemodynamically significant stenoses. A retrospective analysis of 565,504 patients without prior myocardial infarction or revascularization who underwent elective coronary angiography in one of 691 U.S. hospitals revealed that median rate of confirming the presence of CAD was only 45% (interquartile range: 39% to 52%) (3). Beyond this, an additional study showed that within a cohort of 93 patients with 139 vessels in which invasive FFR was measured, the prevalence of a hemodynamically significant stenosis was only 41% (4). Similarly, data from the IRIS-FFR Registry indicated that when FFR was performed on patients with coronary lesions, almost 80% of patients had their revascularization deferred (5). Demographically biased risk scores that use population trends fail consistently in individual patients, especially in populations with no “known” risk factors. A problem of this scale warrants additional methods for the upstream triage of the probability of hemodynamically significant CAD. Currently available methods include exercise ECG and other non-invasive tests such as exercise echocardiography, FFRct, and various nuclear cardiology tests including PET-CT. However, these are expensive and not immediately available. Recently, attention has shifted to the use of artificial intelligence (AI) to assess the inexpensive and widely available 12-lead ECG focusing on detection of left ventricular dysfunction, episodic atrial fibrillation from tracings recorded during normal sinus rhythm, and other structural and valvular diseases (6). We have developed an AI model - ECGio - to predict invasive FFR through the analysis of only a standard 12-lead resting digital ECG, which is a fast, cost-effective, widely accessible, and safe diagnostic method.

## Methods

### Study design

This was a retrospective study involving 1,161 consecutive patients who underwent an elective coronary angiography during which FFR measurements were taken in their coronary vessels at the National Institute of Cardiology in Warsaw, Poland between July 4, 2014 and April 17, 2021. The following groups of patients were excluded: 141 were excluded due to signals with extreme noise or artifact and 8 patients who were duplicates. Of the remaining 1,012 patients, 209 patients who underwent FFR in the left anterior descending artery (LAD) were selected randomly to perform a proof-of-concept cross-validation analysis while holding out the additional 803 patients for a subsequent training exercise and tuning analysis. All patients were 18 years or older. Individuals who were referred for catheterization due to acute coronary syndrome, such as ST-elevation myocardial infarction or non-ST-elevation myocardial infarction, were excluded. Patients with ventricular or atrial rhythm were also excluded due to the presence of a deformed QRS complex. Of the patients analyzed, 53 (25.4%) out of the 209 did not have only LAD disease as defined by either a greatest diameter stenosis <50% in the LAD OR the greatest diameter stenosis ≥50% in the LAD AND a greatest diameter stenosis of ≥50% in the Left Main Artery, Left Circumflex Artery (LCX), or the Right Coronary Artery (RCA). The ECGs of study participants were taken up to and including 7 days prior to angiography; and they were used to train, tune, and test the AI model. ECGs were selected for a five-fold cross-validation training and testing regimen. For each fold, a model was trained on 80% of the FFR measurements and tested on the remaining 20% of unseen FFR measurements; thus, every fold contained either 41 or 42 patients for testing and the remaining 167 or 168 patients for training and tuning (Central Illustration). In each training set, 10% were held out as a tuning set to approximate unseen data. During training, the model that performed best on the tuning set was saved as the representative model for that fold. Performance metrics were then calculated for each fold and the entire database. (**Supplemental Equation 1**).

**Central Illustration.**
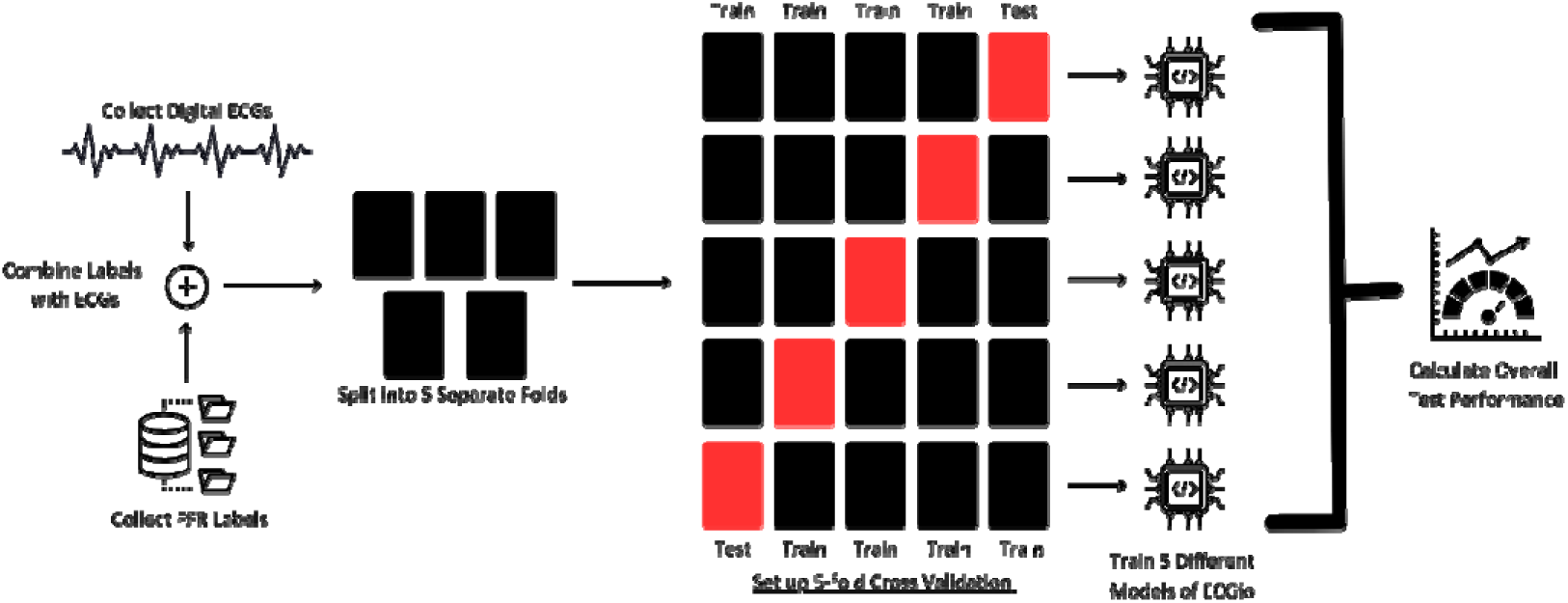
A workflow diagram representing the cross-fold validation process of ECGio. Within the process, the data is split into five folds where each fold uses a different subset of data for training and tuning, the tests on the remainder of the subset.

### Data preparation

The study had two separate datasets -- an ECG signal database and an electronic medical record database with clinical labels, specific coronary angiographic information, and LAD FFR values. The ECG database was composed of de-identified retrospective ECGs extracted via Philips Intellispace. Once the data had been collected and tied to a durable key, the raw digital signal was extracted from the file and converted to time series data and normalized around the x-axis as a baseline point. The electronic medical record database was extracted via query from the center’s electronic medical record to collect each patient’s demographic information, risk factors, and disease information in a deidentified fashion. These two databases were kept separate so as to not bias the model or combine any identifying information.

### AI model – ECGio – and Outcomes

ECGio is a deep learning algorithm built to be used as a cloud-based application for additional ECG interpretation. This study was comprised of both training and testing cycles. Training is the process in which ECGio learns how to correlate the ECG with patient FFR, whereas testing is the process in which model performance is calculated against an independent dataset not seen during training. A brief description of these processes can be seen in **Figure 1**. In this study we analyzed two different outputs for ECGio: 1) a primary analysis as a binary prediction of whether the FFR in the LAD was below a 0.80 threshold and 2) a secondary analysis as a continuous measure for the FFR in the LAD.

**Figure 1.**
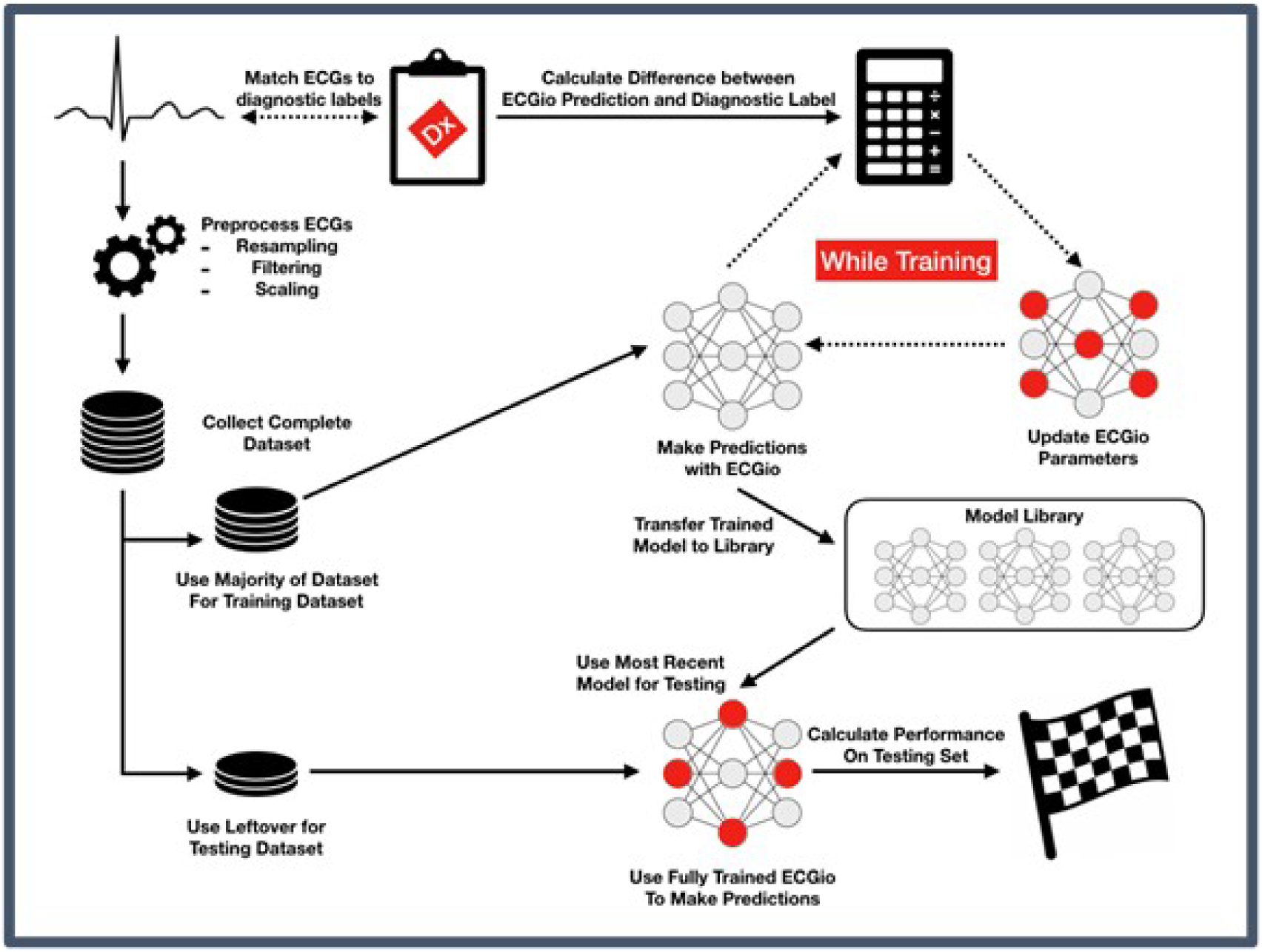
ECGio Product data flow. The workflow for a full training set using ECGio for improvement of the model. In this study, cross- fold validation was employed, so this process was done five times simultaneously with different splits of the data.

The model was trained using a five-fold cross-validation paradigm. Each fold was a randomly selected different group of mutually exclusive patients although there might have been overlap if the population size was not divisible by the number of folds. In the current five-fold cross validation, five different models of ECGio were trained, where four folds were used for training and tuning while one fold was used for testing. The fold used for testing was different in each training period, allowing for a test result on the entire patient population. Performance metrics including sensitivity, specificity, positive predictive value (PPV), negative predictive value (NPV), and F-1 Score were calculated across the five-fold cross-validation for the binary classification. For the continuous measure of FFR, mean average error and root mean squared error were calculated. All calculations were performed using Python 3.7 (Python Software Foundation; Wilmington, DE)

The averaged statistic was calculated with an associated 95% confidence interval. The averaged confidence intervals were calculated using the equations below:

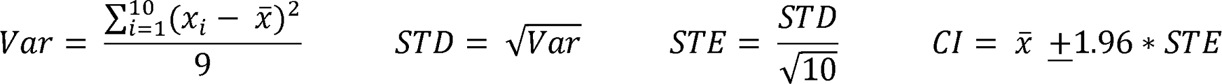

## Results

### Study population

The demographics and baseline clinical data of the 209 patients have been shown in **Table 1**. The prevalence of LAD FFR <0.80 within this study was 65.6% (137/209). Of the 209 patients, 40 (19.1%) had a greatest diameter stenosis ≥50% in another vessel (LCX, RCA, or LM); and 11 (5.3%) had a greatest diameter stenosis ≥70% in another vessel.

**Table 1:**
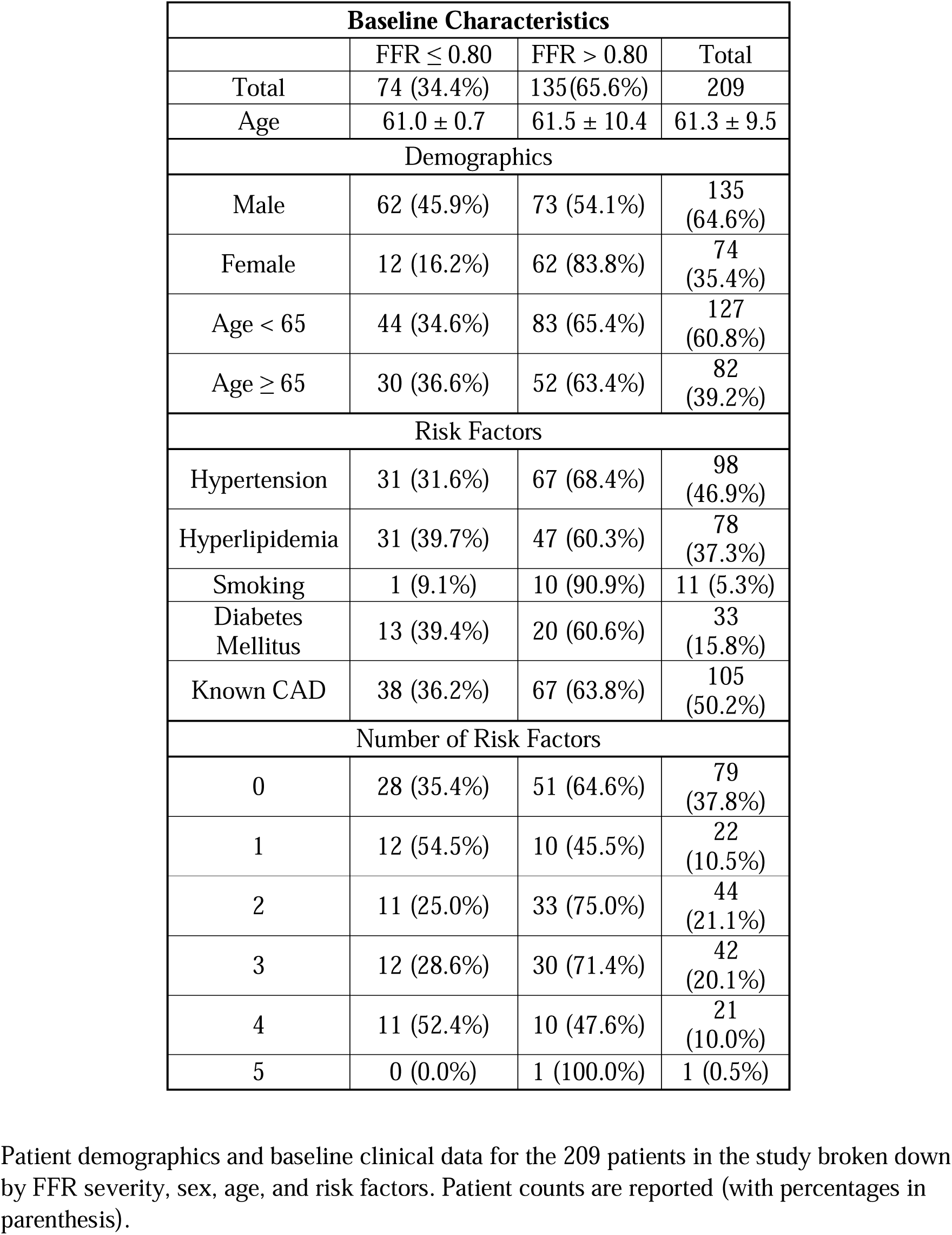
Baseline Patient Demographics.

| Baseline Characteristics |  |  |  |
| --- | --- | --- | --- |
| | FFR ≤ 0.80 | FFR $>$ 0.80 | Total |
| Total | 74 (34.4%) | 135(65.6%) | 209 |
| Age | 61.0 ± 0.7 | 61.5 ± 10.4 | 61.3 ± 9.5 |
| Demographics |  |  |  |
| Male | 62 (45.9%) | 73 (54.1%) | 135<br>(64.6%) |
| Female | 12 (16.2%) | 62 (83.8%) | 74<br>(35.4%) |
| Age $<$ 65 | 44 (34.6%) | 83 (65.4%) | 127<br>(60.8%) |
| Age ≥ 65 | 30 (36.6%) | 52 (63.4%) | 82<br>(39.2%) |
| Risk Factors |  |  |  |
| Hypertension | 31 (31.6%) | 67 (68.4%) | 98<br>(46.9%) |
| Hyperlipidemia | 31 (39.7%) | 47 (60.3%) | 78<br>(37.3%) |
| Smoking | 1 (9.1%) | 10 (90.9%) | 11 (5.3%) |
| Diabetes Mellitus | 13 (39.4%) | 20 (60.6%) | 33<br>(15.8%) |
| Known CAD | 38 (36.2%) | 67 (63.8%) | 105<br>(50.2%) |
| Number of Risk Factors |  |  |  |
| 0 | 28 (35.4%) | 51 (64.6%) | 79<br>(37.8%) |
| 1 | 12 (54.5%) | 10 (45.5%) | 22<br>(10.5%) |
| 2 | 11 (25.0%) | 33 (75.0%) | 44<br>(21.1%) |
| 3 | 12 (28.6%) | 30 (71.4%) | 42<br>(20.1%) |
| 4 | 11 (52.4%) | 10 (47.6%) | 21<br>(10.0%) |
| 5 | 0 (0.0%) | 1 (100.0%) | 1 (0.5%) |

Patient demographics and baseline clinical data for the 209 patients in the study broken down by FFR severity, sex, age, and risk factors. Patient counts are reported (with percentages in parenthesis).

The model’s performance across the five folds has been shown in **Table 2**. The overall ability of ECGio to predict the presence of a reduced FFR this cohort was a sensitivity, specificity, PPV, NPV, accuracy, and F-1 Score of 43.2%, 86.7%, 64.0%, 73.6%, 71.3%, and 51.6%, respectively.

**Table 2:** Cross-fold validation results.

|  | Fold 1 | Fold 2 | Fold 3 | Fold 4 | Fold 5 | **Total** |
| --- | --- | --- | --- | --- | --- | --- |
| True Positives | 9 | 5 | 6 | 5 | 7 | **32** |
| True Negatives | 25 | 22 | 24 | 23 | 23 | **117** |
| False Negatives | 2 | 1 | 7 | 5 | 3 | **42** |
| False Positives | 6 | 14 | 5 | 9 | 8 | **18** |
| Positive Class N | 15 | 19 | 11 | 14 | 15 | **74** |
| Negative Class N | 27 | 23 | 31 | 28 | 26 | **135** |
| Reduced FFR<br>Prevalence | 35.71% | 45.24% | 26.19% | 33.33% | 36.59% | **35.41%** |
| Sensitivity | 0.6 | 0.263 | 0.545 | 0.357 | 0.467 | **0.432** |
| Specificity | 0.926 | 0.957 | 0.774 | 0.821 | 0.885 | **0.867** |
| PPV | 0.818 | 0.833 | 0.462 | 0.5 | 0.7 | **0.64** |
| NPV | 0.806 | 0.611 | 0.828 | 0.719 | 0.742 | **0.736** |
| Diagnostic Accuracy | 0.81 | 0.643 | 0.714 | 0.667 | 0.732 | **0.713** |
| F1-Score | 0.692 | 0.4 | 0.5 | 0.417 | 0.56 | **0.516** |

The model's performance metrics across the five-fold cross-validation. We include a total column that calculates the model's overall performance at the end.

Because age, sex, and risk factors may affect the performance of the model, **Table 3** presents the model’s performance in relation to these factors. In general, there was a slight difference in diagnostic performance between men and women: sensitivity (40.3% in males and 58.3% in females) and specificity (91.8% in males and 80.6% in females). More work is necessary to determine if this was a sex-linked bias or a result of small sample size. In comparison, a smaller difference was observed as a function of age with those 65 years of age and older vs those under age 65 having close diagnostic performance (sensitivity of 40.0% vs 45.5% and specificity of 86.5% vs 86.7%, respectively).

**Table 3:**
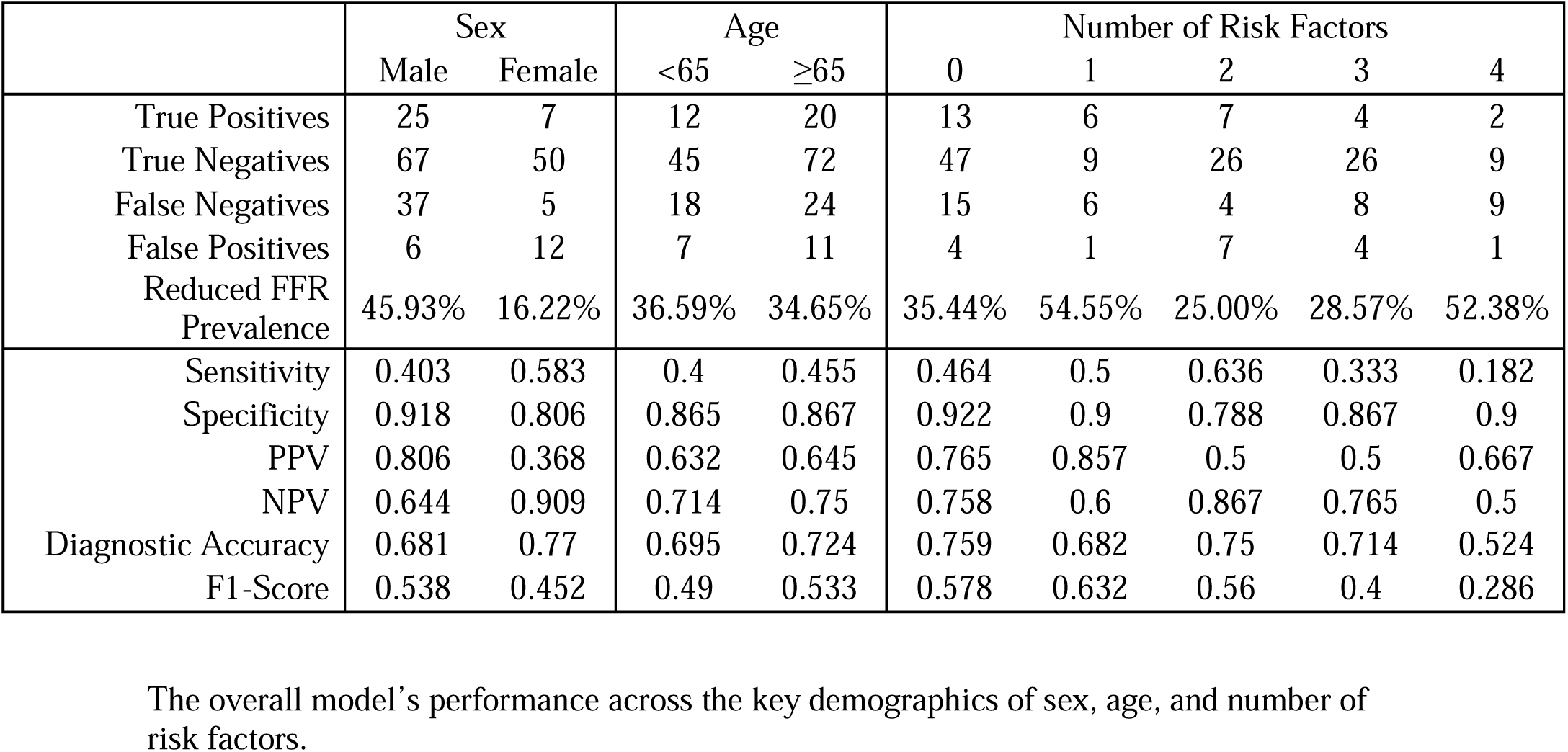
Performance by demographics.

|  | Sex |  | Age |  | Number of Risk Factors |  |  |  |  |
| --- | --- | --- | --- | --- | --- | --- | --- | --- | --- |
|  | Male | Female | <65 | ≥65 | 0 | 1 | 2 | 3 | 4 |
| True Positives | 25 | 7 | 12 | 20 | 13 | 6 | 7 | 4 | 2 |
| True Negatives | 67 | 50 | 45 | 72 | 47 | 9 | 26 | 26 | 9 |
| False Negatives | 37 | 5 | 18 | 24 | 15 | 6 | 4 | 8 | 9 |
| False Positives | 6 | 12 | 7 | 11 | 4 | 1 | 7 | 4 | 1 |
| Reduced FFR<br>Prevalence | 45.93% | 16.22% | 36.59% | 34.65% | 35.44% | 54.55% | 25.00% | 28.57% | 52.38% |
| Sensitivity | 0.403 | 0.583 | 0.4 | 0.455 | 0.464 | 0.5 | 0.636 | 0.333 | 0.182 |
| Specificity | 0.918 | 0.806 | 0.865 | 0.867 | 0.922 | 0.9 | 0.788 | 0.867 | 0.9 |
| PPV | 0.806 | 0.368 | 0.632 | 0.645 | 0.765 | 0.857 | 0.5 | 0.5 | 0.667 |
| NPV | 0.644 | 0.909 | 0.714 | 0.75 | 0.758 | 0.6 | 0.867 | 0.765 | 0.5 |
| Diagnostic Accuracy | 0.681 | 0.77 | 0.695 | 0.724 | 0.759 | 0.682 | 0.75 | 0.714 | 0.524 |
| F1-Score | 0.538 | 0.452 | 0.49 | 0.533 | 0.578 | 0.632 | 0.56 | 0.4 | 0.286 |

The overall model's performance across the key demographics of sex, age, and number of risk factors.

Within the five folds of the cross-validation, ECGio and the gold standard invasive FFR agreed in 148 of the 209 patients, with ECGio tending to underdiagnose stenosis significance. **Figure 2** shows a confusion matrix comparing the frequency of how often ECGio correctly and incorrectly matched the FFR findings across the entire cross-validation. Among the 42 patients in whom ECGio under-predicted stenosis significance, 19 of the patients had FFR values between 0.75 and 0.80 and could be considered borderline.

**Figure 2.**
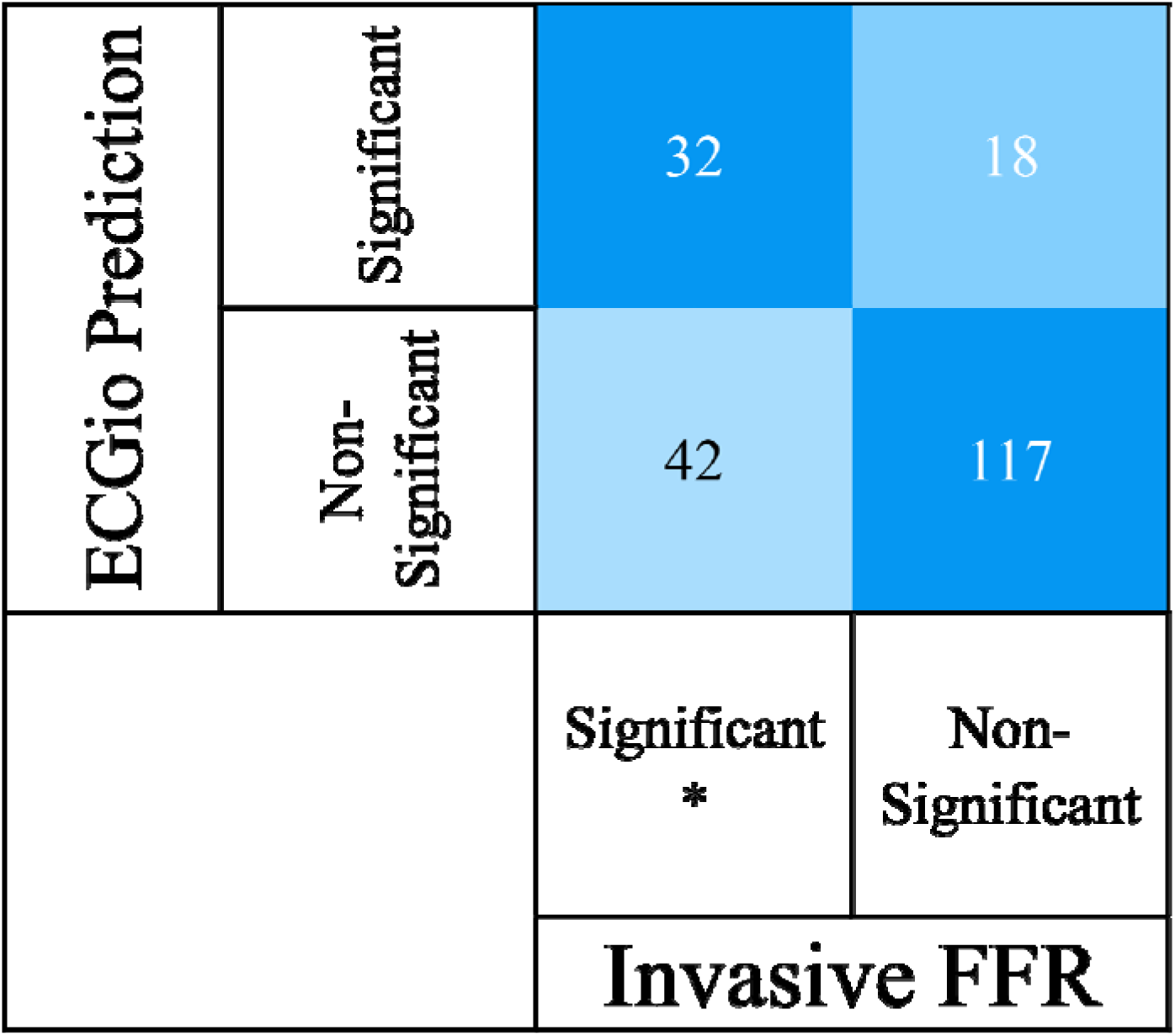
A confusion matrix representing the ECGio prediction of reduced FFR vs invasive reduced FFR. A confusion matrix representing the ECGio prediction of reduced FFR vs the results of the invasive FFR. ECGio represents the rows, where invasive FFR represents the columns.

To see how accurately our model was able to recapitulate FFR values, we looked at three different measures: the mean average error, the root mean squared error, and a Bland-Altman correlation. The mean average error in the patients for measuring the FFR was 1.4%, with the range of FFR values being from 0.39 to 1.00. The Root Mean Squared Error was 7.4%, with a maximum FFR of 1.0, and a minimum FFR of 0.39. The residuals have been visualized in **Figure 3**.

**Figure 3.**
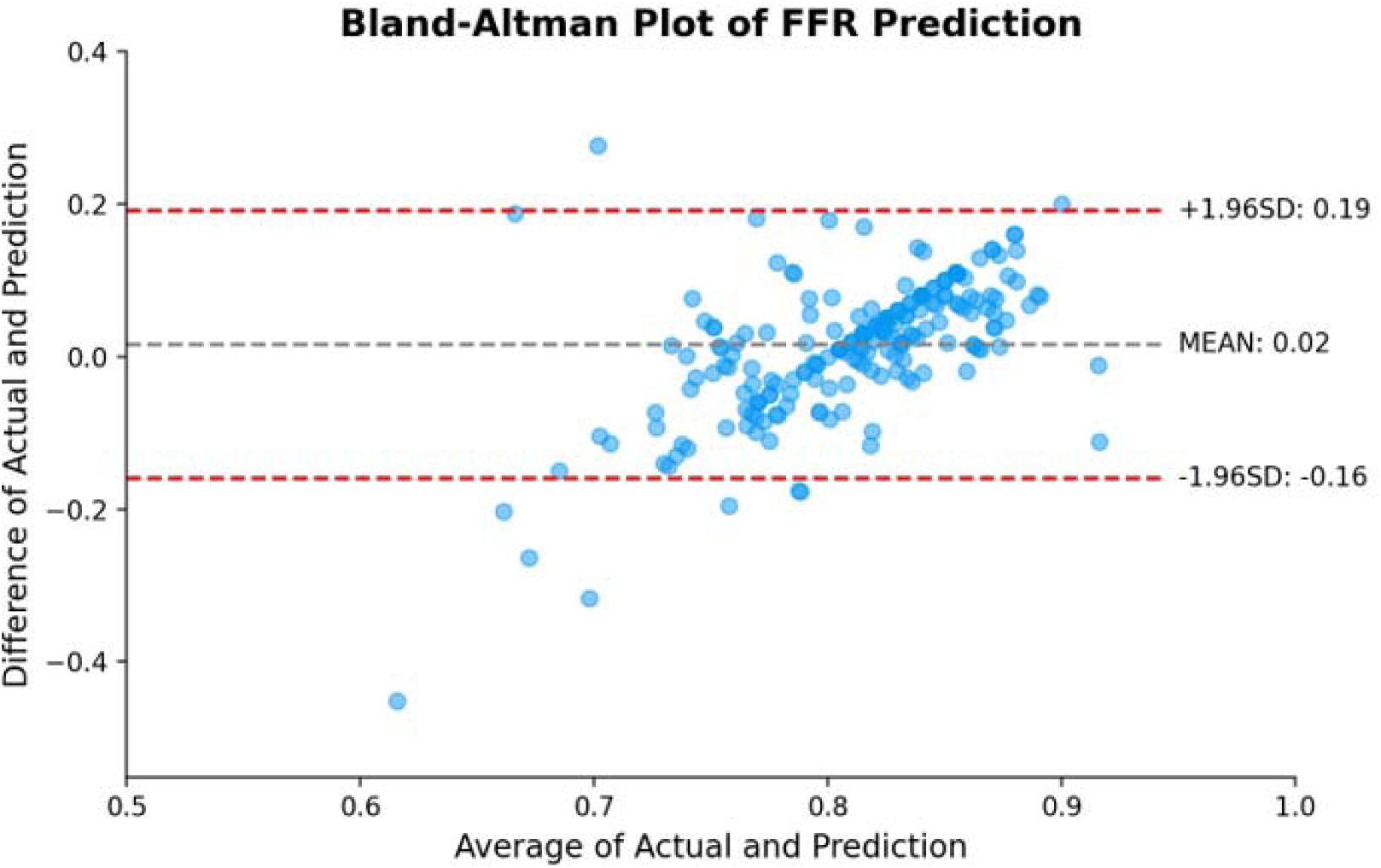
ECGio discrete Bland-Altman plot. A Bland-Altman plot and best fit line for the residuals of the cross-validation results of ECGio in prediction of FFR in comparison against invasive FFR. Red dashed lines represent ± 1.96 SD from the mean.

## Discussion

ECGs are inexpensive, commonly administered tests that measure the cardiac electrical activity by recording voltage difference across the cardiac cycle. For this reason, the use of AI in ECG analysis has recently garnered more attention and is progressing at a rapid pace. Machine learning enables analysis not only of what clinicians can determine from an ECG, but also additional information unable to be discerned by the human eye (6). Machine learning is able to do this by removing a visual processing limitation within a human interpretation, using magnitude values to allow for simultaneous analysis across every point within every ECG lead and their interdependence on one another to create a more complete view of the electromagnetic activity within the heart. However, ECG-based algorithms currently available do not reliably detect the full range of structured cardiac abnormalities much less hemodynamically significant coronary stenosis. Existing rule-based ECG algorithms analyse only a few parts of an ECG, mostly using existing markers and measurements, while ECGio utilizes novel deep learning and AI to evaluate each patient’s likelihood for CAD.

An additional advantage of ECGio is its capability to consistently reconstruct missing or noisy ECG leads, including the reconstruction of a complete 12-lead ECG from just a single lead acquisition device (7). It is extremely useful in devices that, by design, don’t have a full set of 12 leads available.

Previous studies on the diagnosis of CAD via AI-enabled ECG algorithms used the angiographic diameter of stenosis as the standard (8, 9, 10). Unlike this current study, these reports did not provide information about functional severity and the hemodynamic significance of CAD. The current study is the first in which FFR has been determined by AI analysis of a standard resting 12-lead 10-second ECG. Previously, AI has been used to predict this parameter using different methods (6, 11) such as coronary computed tomography angiography (12–15), X-ray coronary angiography (16), optical coherence tomography (17, 18), and intravascular ultrasound (19). The clear advantage of ECG over these techniques is that it is non-invasive, easy to use, inexpensive, real-time, and universally available.

To assess our model’s effectiveness in predicting FFR within the LAD, we referenced similar studies and commercially available tools, as detailed in Table 4 (20). The table compares the performance of the FDA-approved Heartflow FFRct, along with invasive coronary angiography and coronary computed tomography angiography, based on data from their FDA clearance study (NCT01757678). Despite ECGio being trained on a relatively small dataset, it demonstrated competitive results compared to coronary computed tomography angiography and invasive coronary angiography. Given that ECGio is a deep learning-based algorithm, its predictive accuracy is anticipated to improve with the inclusion of more patient data, potentially reaching or surpassing the performance of Heartflow’s FFRct.

**Table 4:** ECGio’s performance metrics in this study versus other studies.

|  | **ECGio** | **CCTA** | **ICA** | **FFRct** |
| --- | --- | --- | --- | --- |
|  | **(this paper)** | **(NXT Trial)** | **(NXT Trial)** | **(NXT Trial)** |
| Patients (N) | 209 | 254 | 254 | 254 |
| Reduced FFR Prevalence | 35.41% | 31.50% | 31.50% | 31.50% |
| Sensitivity | 0.432 (0.365 - 0.499) | 0.938 (0.908 - 0.968) | 0.638 (0.579 - 0.697) | 0.863 (0.821 - 0.905) |
| Specificity | 0.867 (0.821 - 0.913) | 0.339 (0.281 - 0.397) | 0.828 (0.782 - 0.874) | 0.787 (0.737 - 0.837) |
| PPV | 0.640 (0.575 - 0.705) | 0.395 (0.335 - 0.455) | 0.630 (0.571 - 0.689) | 0.651 (0.592 - 0.710) |
| NPV | 0.736 (0.676 - 0.796) | 0.922 (0.889 - 0.955) | 0.832 (0.786 - 0.878) | 0.926 (0.894 - 0.958) |
| Diagnostic Accuracy | 0.713 (0.652 - 0.774) | 0.528 (0.467 - 0.589) | 0.768 (0.716 - 0.820) | 0.811 (0.763 - 0.859) |
| F1-Score | 0.516 (0.448 - 0.584) | 0.556 (0.495 - 0.617) | 0.634 (0.575 - 0.693) | 0.742 (0.688 - 0.796) |
| Invasive | N | N | Y | N |
| Radiation | N | Y | Y | Y |
| Expensive | N | Y | Y | Y |
| Real-Time Analysis | Y | N | N | N |

FFR is a value that is calculated to determine revascularization priority. Lesions with FFR values below 0.75 are worthy of revascularization, while values above 0.80 are considered safe to be deferred. Values in between 0.75 and 0.80 are considered borderline, with treatment decisions up to the discretion of the providing physician. Most invasive angiograms have negative results, and most patients undergoing wire-based FFR have no lesions worthy of revascularization. Non-invasive testing regimes do not accurately filter out patients that do not need invasive therapy (21). This study suggests that, while more training data is needed, ECGio can improve the current diagnostic yield of wire-based FFR. As a non-invasive, widely accessible test, patients with chest pain can be rapidly assessed for needed intervention.

In a recent review, the PRIME checklist proposed comprehensive guidelines for optimizing the study design and data standardization of Cardiovascular Imaging-Related AI models with the goal of eliminating algorithmic errors and biases (22). The adoption of this approach significantly enhances the likelihood of a successful AI application and outlines proper methodologies for clinical studies. ECGio utilizes several of the suggested approaches, with focuses on feature extraction, down sampling of signals, data augmentation, noise removal and normalization, and cross-validation among others.

ECGio has the ability to impact healthcare practise on three primary levels:

- **Reduction in number of false positives sent to catheterization and false negatives released:** ECGio, in early testing, has provided a higher level of accuracy than current standards of care such as stress myocardial perfusion imaging.
- **Reduction in cost of over/undertreatment of chest pain in an emergency setting:** ECGio puts additional diagnostic power in the hands of all medical professionals and allows emergency medical services to better determine treatment pathways for patients in the Emergency Department before they arrive on premise. This will reduce the cost associated with over testing as well as the cost associated with letting a patient go with disease.
- **Increase in quality of preventive care for individual patients through personalization:** By understanding the extent of disease for each patient, physicians will better be able to recommend proper therapeutic life adjustments as well as pharmaceutical intervention when appropriate.

### Study limitations

This research represents a proof-of-concept study; consequently, it has encountered several limitations typical to this category of analysis. The study was limited by a small sample size, particularly for cross-validation resulting in the creation of an AI model. Efficacy of AI is directly correlated to the amount of training data (23). With an expanded sample size, we would likely see stronger performance across all subgroups. The study included only patients who had been referred for angiography. Future studies will analyse patients who undergo invasive and non-invasive FFR entering through other clinical pathways. The study was conducted at a single European site with all white European patients which may result in reduced effectiveness of the algorithm in analysing data from other racial groups (24). However, the scientific society is divided on the significance of such bias. In a recent study (25), the performance of AI designed to detect individuals with left ventricular ejection fraction ≤35% was tested across a range of racial and ethnic subgroups. This convolutional neural network was initially trained on a predominantly homogeneous population of non-Hispanic white individuals (96.2%); yet, it demonstrated similar performance across other subgroups. To maintain a homogeneous dataset in terms of lesion location, we restricted the population to those who obtained FFR in the LAD. This also limited the size of the total dataset. Additional studies are required to examine the performance of ECGio in other lesion locations, but we expect similar performance across lesion locations.

### Conclusions

We trained a novel AI-based algorithm, ECGio, for estimation of invasive FFR in the LAD using only a standard 12-lead resting ECG. The model demonstrated competitive performance to other methods while maintaining an ECG’s non-invasiveness, ease of use, and cost-effectiveness. ECGio has the potential to be employed in healthcare facilities to assist clinicians in screening patients who might require additional diagnostic assessments from a functional perspective. Nevertheless, given the small training and test group, more extensive studies are essential to improve our model.

## Data Availability

All data produced is not available for public use

## Acknowledgement

Not applicable.

## Declaration of Generative AI and AI-assisted technologies in the writing process

During the preparation of this work the authors used chatGPT-3,5 in order to improve readability and language. After using this service, the authors reviewed and edited the content as needed and take full responsibility for the content of the publication.

## Abbreviations

AI: Artificial Intelligence
CAD: Coronary Artery Disease
ECG: Electrocardiogram
FFR: Fractional Flow Reserve
LAD: Left Anterior Descending Artery
LCX: Left Circumflex Artery
LM: Left Main Artery
NPV: Negative Predictive Value
PPV: Positive Predictive Value
RCA: Right Coronary Artery

